# Risk of common psychiatric disorders, suicidal behaviours and premature mortality following violent victimisation: A matched cohort and sibling-comparison study of 127,628 people who experienced violence in Finland and Sweden

**DOI:** 10.1101/2024.05.08.24307040

**Authors:** Amir Sariaslan, Joonas Pitkänen, Jonas Forsman, Ralf Kuja-Halkola, Isabell Brikell, Brian M. D’Onofrio, Mikko Aaltonen, Henrik Larsson, Pekka Martikainen, Paul Lichtenstein, Seena Fazel

**Author notes:** **All correspondence should be sent to**: Dr Amir Sariaslan, Department of Psychiatry, University of Oxford, Warneford Hospital, Oxford OX3 7JX, United Kingdom.

## Abstract

**Background:** Associations between violent victimisation and psychiatric disorders are hypothesised to be bidirectional, but the role of violent victimisation in the aetiologies of psychiatric disorders and other adverse outcomes remains unclear. We aimed to estimate associations between violent victimisation and subsequent common psychiatric disorders, suicidal behaviours, and premature mortality whilst accounting for unmeasured familial confounders.

**Methods and Findings:** Using nationwide registers, we identified a total of 127,628 individuals born in Finland (1987-2004) and Sweden (1973-2004) who had experienced violent victimisation, defined as either hospital admissions or secondary care outpatient visits for assault-related injuries. These were age- and sex-matched with up to 10 individuals in the general population (n=1,276,215) and their unaffected siblings (n=132,408). Outcomes included depression, anxiety, personality disorders, substance use disorders, suicidal behaviours, and premature mortality. Participants were followed from the victimisation date until the date of the outcome, emigration, death, or December 31, 2020, whichever occurred first. Country-specific associations were estimated using stratified Cox regression models, which also accounted for unmeasured familial confounders via sibling comparisons. The country-specific associations were then pooled using meta-analytic models.

Among 127,628 patients (69.0% men) who had experienced violent victimisation, the median age at first violent victimisation was 21 (interquartile range: 18-26) years. Absolute risks of all outcomes were larger in those who were exposed to violent victimisation compared to population controls (2.3-22.5 vs. 0.6-7.3 per 1000 person-years). In adjusted models, people who had experienced violent victimisation were between two to three times as likely as their siblings to develop any of the outcomes (adjusted hazard ratios [aHRs]: 1.7-3.0). Risks remained elevated two years post-victimisation (aHRs: 1.4-2.3).

**Conclusions:** Improving clinical assessment, management and aftercare psychosocial support could potentially reduce rates of common psychiatric disorders, suicidality and premature in individuals experiencing violent victimisation.

---

People diagnosed with psychiatric disorders are at least twice as likely as the general population to be violently victimised (1–3). This finding has been consistently replicated across several victimisation outcomes, including self-reports (1), police reported events (4,5), assaults requiring medical care (6,7), and homicidal deaths (8). Although it is commonly hypothesised that associations between common psychiatric disorders and violent victimisation are bidirectional in nature, few studies have rigorously examined whether exposure to violent victimisation contributes to the development of common psychiatric disorders and suicidal behaviours.

Two key methodological limitations are present in previous work that has considered violent victimisation as a risk factor for psychiatric disorders. First, violent victimisation has been combined with several broader indices of adverse childhood experiences, bullying or negative/stressful life events (2,9,10). While this has the benefit of simultaneously considering multiple types of adversities, they do not permit the evaluation of the specific contributions of severe violent victimisation to aetiologies of psychiatric disorders, suicidal behaviours, and premature mortality.

Second, twin studies have consistently demonstrated that victimisation is moderately heritable, with genetic factors accounting for around 20-40% of the variance (11,12). These findings are expected, as victimisation is also associated with a vast array of risk factors that are at least moderately heritable, including low cognitive abilities (13), personality traits (14), and violent perpetration (15). As these risk factors are implicated in the aetiologies of common psychiatric disorders, suicidal behaviours, and premature mortality, unmeasured genetic confounding could account for at least a portion of the associations. The few genetically informative studies that have examined the long-term associations between violent victimisation and subsequent common psychiatric disorders have reported conflicting findings, in part due to limited statistical power.

A recent study (16) that considered unmeasured familial confounding in Sweden reported that people who had experienced an assault were three times as likely as their unaffected siblings to develop any psychiatric disorders within the year following the assault, and twice as likely thereafter. However, follow-up of the study ended in 2013, many relevant victimisation exposures (i.e., assaults by strangulation and poisoning) were not included, sex-specific associations were not investigated for specific outcomes, and findings have not been externally replicated.

To address these evidence gaps, we examined the extent to which exposure to severe forms of violent victimisation was associated with subsequent risks of common psychiatric disorders, suicidal behaviours, and premature mortality by pooling nationwide register data from Finland and Sweden. In complementary analyses, we also tested for effect moderation by pre-existing psychiatric disorders. We were further able to account for unmeasured familial confounders, such as shared genetic and early environmental risk factors, by comparing risks of the outcomes between siblings who were differentially exposed to violent victimisation. We complemented the sibling-comparison design by additionally accounting for measured confounders that varied between the siblings over time, such as being exposed to single motherhood and low family income at birth.

## Methods

### National Registers

A unique personal identification number is assigned either at birth or upon immigration to all Finnish and Swedish residents, thus enabling accurate linkage across various national social and health registers (17,18). We were granted access to pseudonymised administrative population data by the Ethics Board of Statistics Finland (TK-53-1490-18), the Finnish Institute of Health and Welfare (THL/2180/14.02.00/2020), and the Swedish Ethical Review Authority (Dnr 2020-06540; Dnr 2022-06204-02). Informed consent is neither mandated by Finnish nor Swedish law for research involving national registers with pseudonymised identifiers. The study followed the STROBE reporting guidelines (**eTable 1**), but did not have a prospective analysis plan.

The Care Register for Health Care, maintained by the Finnish Institute for Health and Welfare (THL), and the National Patient Register, maintained by the Swedish National Board for Health and Welfare, contain information on inpatient hospitalisation episodes (Finland: 1969-2020, Sweden: 1973-2020) and specialist outpatient care (Finland: 1998-2020; Sweden: 2001-2020). Diagnoses were classified according to the Finnish and Swedish versions of the International Classification of Diseases (ICD) from the eighth to tenth revisions. Data on mortality dates, including the primary and contributing causes of death, were extracted from the Causes of Death Registers of each country, which utilised the same ICD classifications as the hospital data. Data on sociodemographic factors, including emigration dates, were retrieved from a series of population administrative registers maintained by Statistics Finland and Statistics Sweden.

Using these registers, we identified 1,090,641 individuals born in Finland between 1987 and 2004, and 3,269,545 individuals born in Sweden between 1973 and 2004. Individuals who could not be linked to their biological parents (n_Finland_=15,406; n_Sweden_=42,340) or who lacked data on parental socio-demographic factors (n_Finland_=1900; n_Sweden_=31,103) were excluded. The resulting analytical samples comprised 98.4% (n=1,073,335) of the targeted Finnish sample and 95.9% (n= 3,196,102) of the Swedish sample, yielding a pooled sample of 4,269,437 individuals.

### Violent victimisation

Violent victimisation was defined as an inpatient care episode or secondary care outpatient visit associated with a diagnosis of an injury purposefully inflicted by another person (ICD-codes are provided in **eTable 2**; diagnostic validity of the examined measures is discussed in **eText 1**).

### Common psychiatric disorder, suicidal behaviours, and premature mortality

The following common psychiatric disorders were investigated as outcomes: depression, anxiety, personality disorders, alcohol use disorders, and substance use disorders. The measures of alcohol and substance use disorders also included deaths attributable to these conditions. We additionally investigated suicidal behaviours, encompassing both non-fatal self-harm and completed suicide, as well as premature mortality, defined consistent with the literature as any death prior to the age of 65 years (19). We further included pre-existing measures of the psychiatric disorders and self-harm.

### Control groups

We matched each index person experiencing violent victimisation with up to ten individuals from the general population without such experiences throughout the follow-up by sex and birth year. These control individuals were required to be alive and residing in Finland or Sweden at the time of the first violent victimisation event of the index person. We also matched each index person with their biological full-siblings who had not experienced violent victimisation to account for unmeasured shared familial confounding. In these analyses, we included all siblings and adjusted for sex and birth year differences using statistical covariates. If multiple siblings in a given family had experienced violent victimisation, we matched them with all their unaffected co-siblings in separate strata (19). To account for the fact that the same individuals could serve as controls across multiple strata, we specified all sibling models to estimate individual-level cluster-robust standard errors.

By comparing outcome rates between differentially exposed siblings, we accounted for time invariant and unmeasured familial confounders shared between the siblings, including an average of half of their co-segregating genes and their shared childhood environments (20). The extent to which the sibling comparisons were attenuated from the population estimates indicated the influence of these unmeasured familial confounders on the studied associations, assuming no misclassification of the exposure and outcome variables.

The starting points for both index persons and control groups were defined as the date of the first violent victimisation event. Participants were censored if they migrated, died, experienced the outcome of interest, or reached the endpoint of the study on December 31, 2020.

### Analytical strategy

In each country, we fitted separate stratified Cox proportional hazards analyses to estimate adjusted hazard ratios (aHRs) for the risk of common psychiatric disorders, suicidal behaviours, and premature mortality among individuals who had experienced violent victimisation compared with matched individuals without such an experience, with time in years since the first victimisation event (or equivalent date for the matched individuals) as the underlying time scale. As each index person who experienced violent victimisation and their corresponding control individuals were assigned to a separate stratum, the models estimated varying baseline hazards across these strata, indicating that comparisons were made within each stratum.

We fitted three models that gradually accounted for a larger set of potential confounders. The ‘Crude’ model only accounted for the matched characteristics (e.g., sex and birth year). The ‘Adjusted’ model, additionally accounted for the following measured confounders (definitions in **eTable 3**): birth order, immigrant background, parental characteristics measured at birth (low family income, single mother, parental psychiatric disorders, and violent criminality) in addition to any psychiatric disorders and self-harm that had occurred prior to the victimisation event (or the equivalent period for the controls). The ‘Sibling-comparison’ model further accounted for unmeasured familial confounders by re-running the adjusted model on a sample that included index persons who had experienced violent victimisation and their biological full-siblings who did not.

To assess the moderating role of pre-existing psychiatric disorders and self-harm behaviours on the associations under study, we added an interaction term to the sibling-comparison models. We used an aggregated binary measure of all pre-existing conditions to reduce the number of hypothesis tests. To aid interpretation, we estimated linear combinations of the model parameters to assess the differential associations of pre-existing conditions in the presence and absence of violent victimisation. Interaction terms that did not reach statistical significance (P>0.05) were omitted from the models.

Consistent with prior studies (21,22), we fitted inverse variance-weighted fixed-effects meta-analysis models to the resulting regression coefficients and standard errors from the country-specific models (23), which allowed us to pool the associations across both countries by weighting the magnitude of the associations by their sample sizes.

In complementary sensitivity analyses, we examined an alternative exposure measure, which included individuals who were registered as victims of violent offences (e.g., attempted homicide, assault, robbery, unlawful threats, or sexual offences) in Finnish police reports between 2008 and 2020 and who did not have corresponding hospital episode. Additional sensitivity analyses are described in **eText 2**.

## Results

A total of 127,628 individuals had experienced violent victimisation in Finland and Sweden at a median age of 21 years, with men accounting for 69% of the group (**Table 1; eTables 4-5**). When compared to age and sex-matched controls (n=1,276,215), those who experienced violent victimisation were more likely to come from backgrounds characterised by low family income, a single mother household, and parents with a history of psychiatric disorders and violent crime convictions at offspring birth (**Table 1**).

**Table 1.** Baseline demographic characteristics.

|  | General population controls | Individuals exposed to violent victimisation | Unaffected sibling controls | Individuals exposed to violent victimisation with unaffected siblings |
| --- | --- | --- | --- | --- |
| **Total** | 1,276,215 | 127,628 | 132,408 | 85,825 |
| **Median age at victimisation (IQR) years** | - | 21 (18-26) | - | 21 (18-25) |
| **Sex** |  |  |  |  |
| Female | 395,990 (31.0%) | 39,599 (31.0%) | 66,048 (49.9%) | 25,714 (30.0%) |
| Male | 880,225 (69.0%) | 88,029 (69.0%) | 66,360 (50.1%) | 60,111 (70.0%) |
| **Birth cohorts** |  |  |  |  |
| 1973-1974 | 51,940 (4.1%) | 5194 (4.1%) | 3446 (2.6%) | 1990 (2.3%) |
| 1975-1979 | 134,290 (10.5%) | 13,429 (10.5%) | 13,021 (9.8%) | 7820 (9.1%) |
| 1980-1984 | 179,270 (14.0%) | 17,927 (14.0%) | 19,316 (14.6%) | 12,495 (14.6%) |
| 1985-1989 | 307,062 (24.1%) | 30,712 (24.1%) | 29,948 (22.6%) | 21,530 (25.1%) |
| 1990-1994 | 327,253 (25.6%) | 32,726 (25.6%) | 34,519 (26.1%) | 23,700 (27.6%) |
| 1995-1999 | 184,320 (14.4%) | 18,432 (14.4%) | 20,740 (15.7%) | 13,070 (15.2%) |
| 2000-2004 | 92,080 (7.2%) | 9208 (7.2%) | 11,418 (8.6%) | 5220 (6.1%) |
| **Birth order** |  |  |  |  |
| 1st | 773,997 (60.6%) | 75,172 (58.9%) | 65,569 (49.5%) | 47,213 (55.0%) |
| 2nd | 365,732 (28.7%) | 36,619 (28.7%) | 40,939 (30.9%) | 26,881 (31.3%) |
| 3rd | 101,845 (8.0%) | 11,537 (9.0%) | 16,676 (12.6%) | 8456 (9.9%) |
| 4th or higher | 34,641 (2.7%) | 4300 (3.4%) | 9224 (7.0%) | 3275 (3.8%) |
| **Immigration background** | 197,632 (15.5%) | 26,495 (20.8%) | 26,658 (20.1%) | 16,571 (19.3%) |
| **Mother single at offspring birth** | 520,460 (40.8%) | 63,189 (49.5%) | 53,410 (40.3%) | 37,770 (44.0%) |
| **Family income in the bottom decile at offspring birth** | 119,059 (9.3%) | 17,018 (13.3%) | 13,165 (9.9%) | 9332 (10.9%) |
| **Parental history of psychiatric disorders** | 54,400 (4.3%) | 11,381 (8.9%) | 9594 (7.2%) | 6048 (7.0%) |
| **Parental history of violent criminality** | 67,502 (5.3%) | 16,476 (12.9%) | 14,644 (11.1%) | 9439 (11.0%) |
| **Pre-victimisation psychiatric disorders and self-harm** |  |  |  |  |
| Depression | 42,081 (3.3%) | 13,327 (10.4%) | 6833 (5.2%) | 8300 (9.7%) |
| Anxiety | 45,993 (3.6%) | 14,679 (11.5%) | 7113 (5.4%) | 9009 (10.5%) |
| Personality disorder | 7799 (0.6%) | 4419 (3.5%) | 1482 (1.1%) | 2582 (3.0%) |
| Alcohol use disorder | 23,728 (1.9%) | 12,125 (9.5%) | 4346 (3.3%) | 7458 (8.7%) |
| Drug use disorder | 10,550 (0.8%) | 9497 (7.4%) | 2268 (1.7%) | 5502 (6.4%) |
| Other psychiatric disorders | 68,705 (5.4%) | 11,928 (9.3%) | 8730 (6.6%) | 7494 (8.7%) |
| Self-harm | 23,304 (1.8%) | 10,483 (8.2%) | 4095 (3.1%) | 6382 (7.4%) |

The median follow-up time was 9.5 years across all outcomes (**Table 2; eTable 6**). Individuals who had experienced violent victimisation had considerably elevated rates of all outcomes compared to the population controls (range: 2.3-22.5 vs. 0.6-7.3 per 1000 person-years; Table 2). Individuals who had experienced violent victimisation were at least three times as likely as the population controls, matched for sex and birth year, to have experienced any of the outcomes (**Figure 1**), ranging from a three-fold elevated risk for being diagnosed with depression (adjusted hazard ratio [aHR]=2.9; 95% CI: 2.8-2.9) to over an eight-fold risk increase of being diagnosed with a drug use disorder (aHR=8.3; 95% CI: 8.2-8.5). These estimates were attenuated as we gradually accounted for measured confounders (aHRs: 2.0-4.8) and unmeasured familial confounders (aHRs: 1.7-3.0).

**Figure 1.**
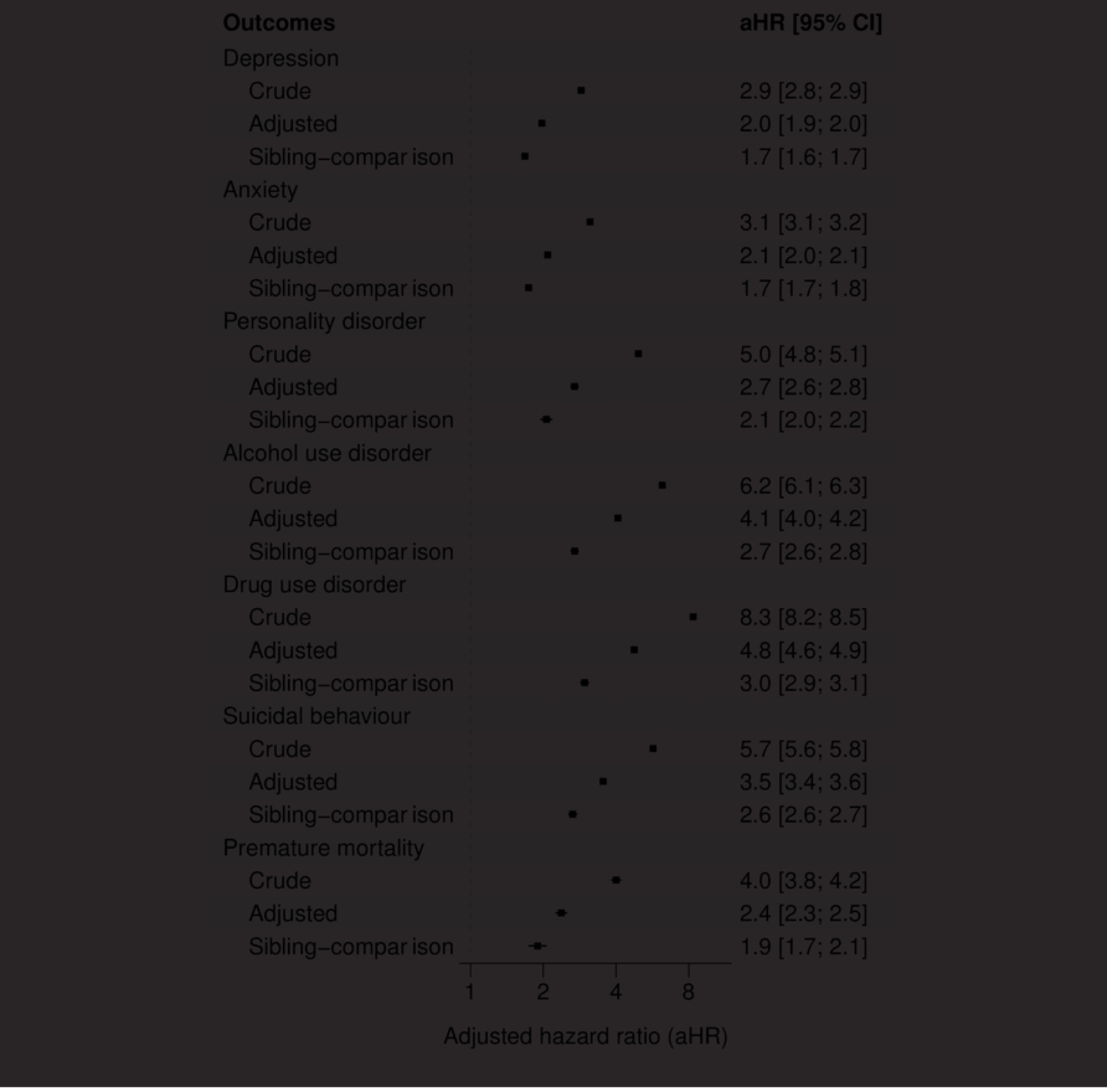
Population-wide and within-family associations between violent victimisation and subsequent common psychiatric disorders, suicidal behaviours, and premature mortality in Finland and Sweden. Notes: The ‘Crude’ model adjusted for sex and birth year. The ‘Adjusted’ model additionally accounted for birth order, parental immigrant background, low family income at offspring birth, single mother at offspring birth, parental psychiatric and violent crime histories at offspring birth, as well as pre-victimisation psychiatric disorders, substance use disorders and self-harm. The ‘Sibling-comparison’ model refers to within-family estimates comparing biological full-siblings differentially exposed to violent victimisation and is adjusted for all time-invariant unmeasured familial confounders shared between the siblings as well as the following measured confounders that vary within families: sex, birth year, birth order, and parental characteristics at birth (low family income, single mother, psychiatric history, and violent crime history) as well as any pre-existing psychiatric disorders and self-harm events.

**Table 2.**
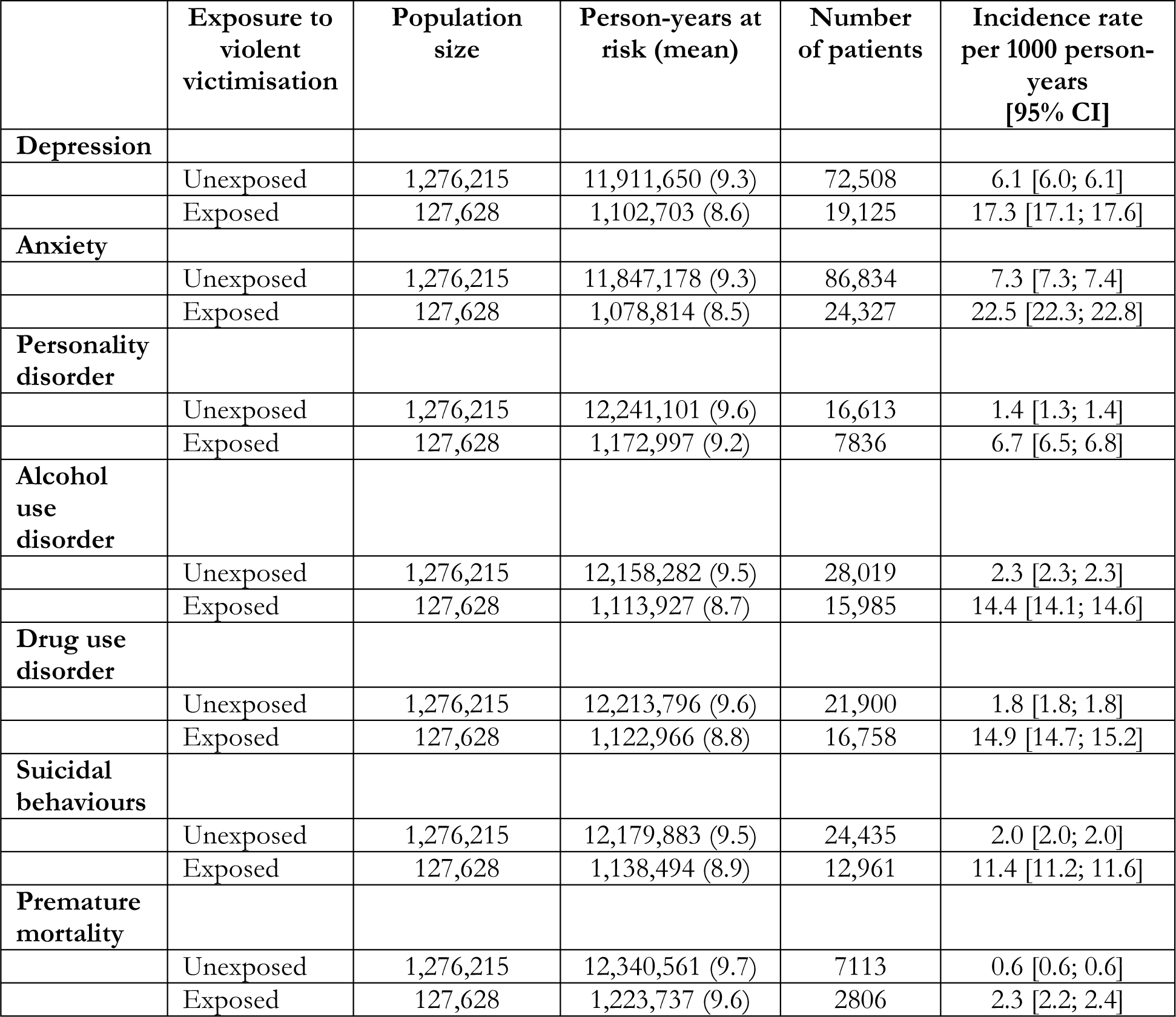
Person-time at risk, number of individuals with the outcomes, and incident rates per 1000 person-years for common psychiatric disorders, suicidal behaviours, and premature mortality stratified across individuals exposed to violent victimization.

There was little evidence the presented associations varied systematically across the included countries, birth cohorts or age at victimisation categories (**eFigures 1-3**). However, the association between violent victimisation and suicidal behaviours was more pronounced in Finland than in Sweden (aHRs: 3.5 vs 2.6; **eFigure 1**). Furthermore, the associations tended to be stronger among sisters than brothers (aHRs: 1.8-3.8 vs. 1.6-2.5; **eFigure 4**) across all outcomes except for premature mortality (aHRs: 1.7-1.7; **eFigure 4**).

We found some evidence in support of a moderating effect of pre-existing psychiatric disorders and self-harm (**eTable 7**). For depression, however, this effect modification appeared to be negative, which suggests that the combined contributions of pre-existing conditions and violent victimisation were slightly weaker than the sum of their individual contributions (aHR_interaction term_: 0.9). Conversely, for alcohol and drug use disorders as well as suicidal behaviours, the effect modification was positive (aHRs_interaction terms_: 1.2-1.3), indicating that the joint contributions of pre-existing conditions and violent victimisation appeared to be greater than the sum of their individual contributions. By examining the four exposure groups (i.e., siblings with and without pre-existing psychiatric disorders, self-harm, and violent victimisation experiences), we found negligible differences from the primary findings when restricting the analyses to those without pre-existing psychiatric disorders and self-harm episodes (aHRs: 1.8-2.9 vs. 1.7-3.0; **Figures 1-2**). Among those with pre-existing conditions, however, we found that violent victimisation was associated with considerably higher rates of all outcomes (aHRs: 4.9-12.5 vs. 2.4-3.6; **Figure 2**).

**Figure 2.**
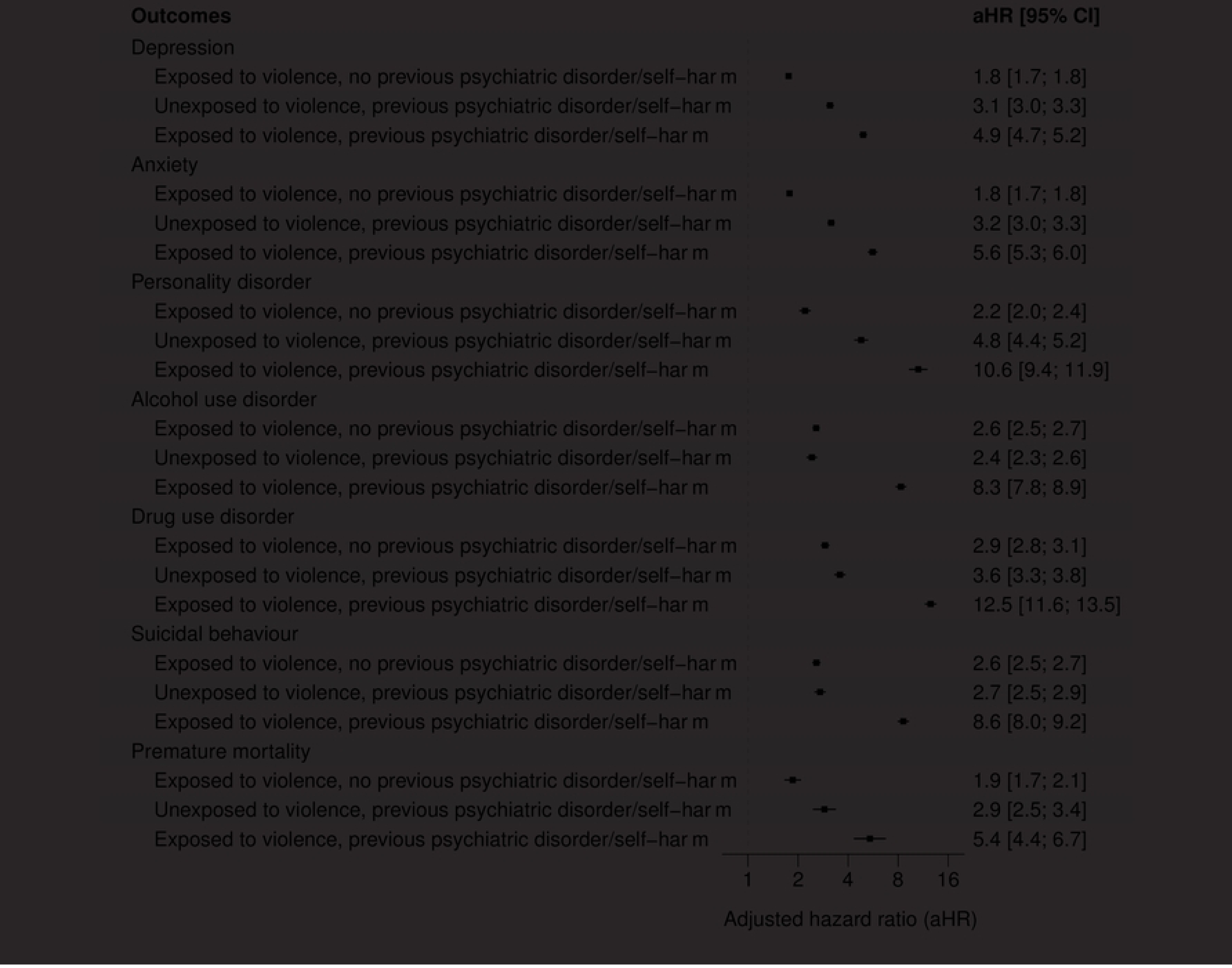
Within-family associations between violent victimisation and subsequent common psychiatric disorders, suicidal behaviours, and premature mortality among Finnish and Swedish siblings with and without previous psychiatric disorders or self-harm. Notes: The reference group included co-siblings who were unexposed to violence and had no pre-existing psychiatric disorders or self-harm events. The estimates refer to within-family associations comparing biological full-siblings differentially exposed to violent victimisation and is adjusted for all time-invariant unmeasured familial confounders shared between the siblings as well as the following measured confounders that vary within families: sex, birth year, birth order, and parental characteristics at birth (low family income, single mother, psychiatric history, and violent crime history) as well as any pre-existing psychiatric disorders and self-harm events.

In Finland, where we additionally investigated a different method to ascertain victimisation, associations between hospital or police recorded victimisation, and outcomes were similar in magnitude (**Figure 3**). The only exception involved suicidal behaviours; assaults requiring medical attention were associated with elevated risks than assaults that had only been reported to the police (aHR=3.5 vs. 2.6). Starting the follow-up period up to two years after the first victimisation date had a marginal impact on the sibling-comparison estimates across both countries, thus suggesting that the associations remained over a longer time period (aHRs: 1.4-2.3; **eFigure 5**). We found commensurate results when adjusting for additional indicators of low parental socioeconomic status (**eFigure 6**), requiring a minimum of two diagnoses of the psychiatric disorders on separate occasions (**eFigure 7**), and considering victimisation events that occurred after 2005 (**eFigure 8**).

**Figure 3.**
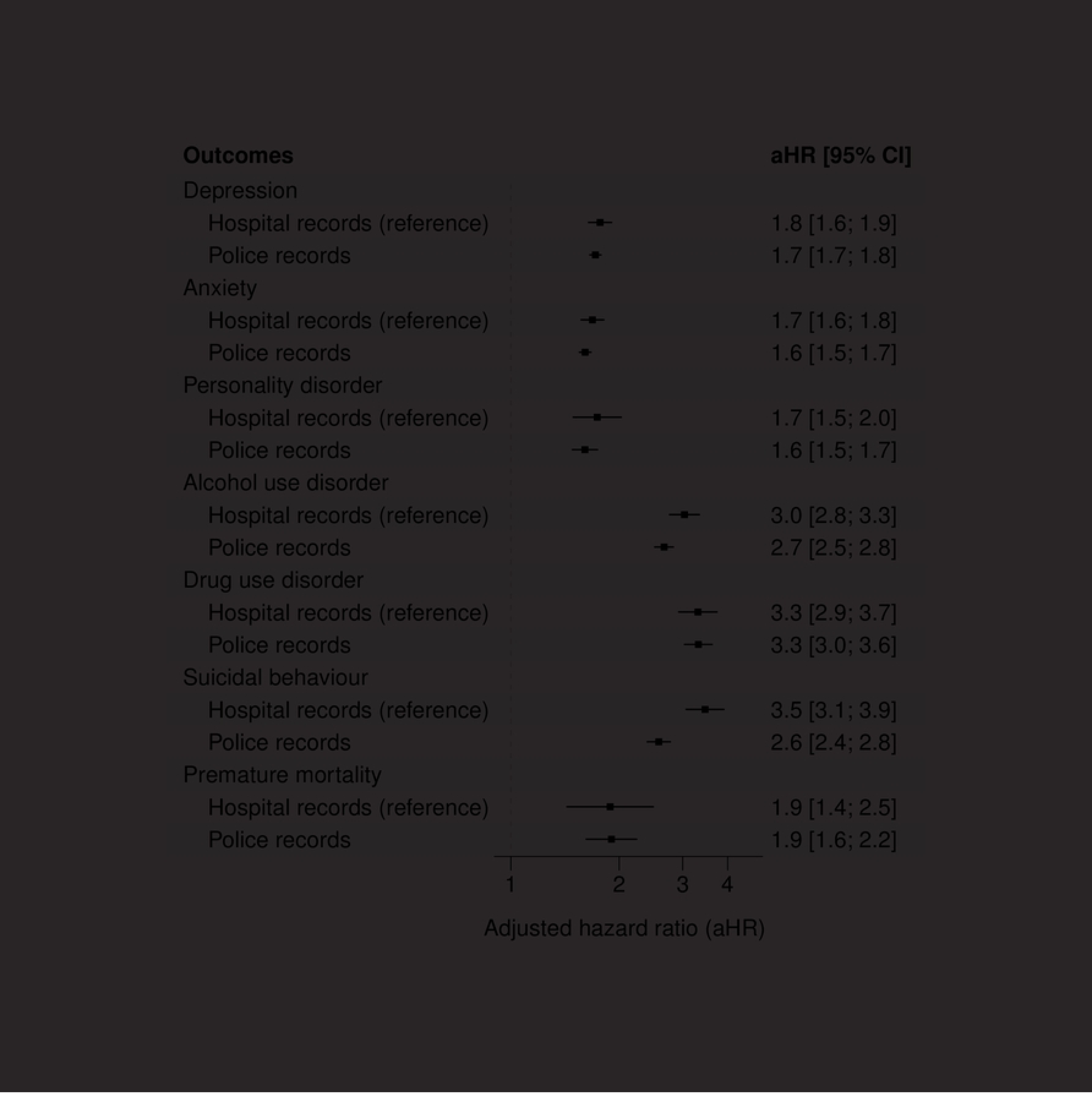
Within-family associations between specific types of violent victimisation (e.g., hospital or police records) and subsequent common psychiatric disorders, suicidal behaviours, and premature mortality among Finnish siblings. Notes: The analyses of police recorded violent victimisation were based on 43,487 Finnish individuals born between 1987 and 2004 who were registered by law enforcement as being victims of violent crimes but had not received specialised medical care for their injuries. They were matched with their unexposed siblings born during the same period (n=68,107). For improved comparability, the reference estimates for hospital records were solely based on Finnish siblings. The estimates refer to within-family associations comparing biological full-siblings differentially exposed to violent victimisation and is adjusted for all time-invariant unmeasured familial confounders shared between the siblings as well as the following measured confounders that vary within families: sex, birth year, birth order, and parental characteristics at birth (low family income, single mother, psychiatric history, and violent crime history) as well as any pre-existing psychiatric disorders and self-harm events.

## Discussion

In this cross-national cohort study of 127,628 individuals who were exposed to violent victimisation, each matched with up to 10 general population controls of the same age and sex, we investigated associations between violent victimisation and subsequent risks of common psychiatric disorders, suicidal behaviours, and premature death. We report four key findings.

First, we found that people who experienced violent victimisation had considerably elevated rates of common psychiatric disorders, suicidal behaviours and premature mortality compared to age and sex matched individuals in the general population (median rates: 17.3 vs 6.1 events per 1000 person-years). Expressed as relative risks, those who had experienced violent victimisation had at least a three-fold increased risk of the examined outcomes compared to the general population controls. Following more rigorous adjustments for an extensive set of measured confounders that captured sociodemographic background factors and pre-existing diagnoses of psychiatric disorders and self-harm, the associations were attenuated but remained strong. These findings are consistent with studies that have reported elevated rates of psychiatric disorders and related behavioural outcomes in people who have been exposed to various measures of victimisation, including peer victimisation (24,25,10) and intimate partner violence (26–28).

Second, our estimates were further attenuated once we had accounted for unmeasured familial confounders, but they still indicated that violent victimisation was associated with least a two-fold elevated risk of all outcomes. We tested for unmeasured familial confounding by comparing outcome rates in people who had experienced violent victimisation with their siblings without such experiences. This approach was especially informative for the associations with alcohol use disorders, drug use disorders and suicidal behaviours, which were attenuated from a four to five-fold risk increase in the adjusted population-wide models to a three-fold risk increase in the sibling-comparison models. These findings add to previous work that has shown the importance of accounting for unmeasured familial confounders when assessing mental health outcomes following violent victimisation (16,29,30). In addition, it suggests that previously reported associations (31) without such adjustments are likely overestimated.

Third, individuals with a history of psychiatric disorders and self-harm, but no experience of violent victimisation, had a three to four-fold increased risk of experiencing the examined outcomes compared to their siblings lacking such experiences. However, those who had experienced violent victimisation, in addition to having pre-existing conditions, were around 5-10 times as likely as their siblings without any psychiatric or victimisation history to experience the outcomes. These findings suggest that the bidirectional relationship (6,5,16) between violent victimisation and psychiatric disorders and related behaviours may potentially follow an amplification model in which the exposures and outcomes mutually reinforce each other over time.

Fourth, we were able to replicate our findings by using a different objective method for measuring violent victimisation, which was police-reported violent victimisation. This approach allowed us to identify over twice as many people experiencing violent victimisation, and this replication suggests that our main findings may be generalisable to violent victimisation that does not lead to medical attention.

Family-based studies that have examined associations between violent victimisation and subsequent risks for psychiatric disorders and related behavioural outcomes have reported conflicting findings to date (29,30). Our findings are nevertheless consistent with a Swedish sibling-comparison study (16) with follow-up that ended in 2013. In addition to updated data and external replication, the present study examined several important violent victimisation exposures that were not investigated in the prior study, including assaults by strangulation and poisoning, as well as sex-specific associations between violent victimisation and a range of psychiatric disorders and suicidal behaviours. Moreover, our findings are also consistent with a recent within-individual study (22) conducted using nationwide registers that reported a 2-3-fold increased odds of self-harm occurring in the week following violent victimisation compared to earlier periods within the same person. The present study uses a more comprehensive measure of violent victimisation and investigates longer-term impact on multiple psychiatric outcomes. We also investigated premature mortality, a novel outcome of increasing importance to public mental health.

Strengths include the use of high-quality nationwide register data with over 127,000 people who had been assaulted to the extent that they required specialist medical care, and around 43,500 individuals who had been identified by the police as victims of a violent offence. The registers allowed us to match each of these individuals with general population controls of the same age and sex, and also with their siblings without violent victimisation experiences to rigorously account for their shared and time-invariant unmeasured familial confounders.

There are some limitations. First, our approach to measuring violent victimisation, common psychiatric disorders, and suicidal behaviours was based on diagnosis with these conditions and injuries in secondary care settings, reflecting the more severe presentation of these. Moreover, outpatient care data was unavailable before 1998 in Finland and 2001 in Sweden. While alternative methods, such as self-reporting and proxy-reporting, can be employed to assess less severe forms of violent victimisation and psychiatric disorders, they have important methodological limitations, including non-response, reporting biases of sensitive information and longitudinal attrition. Moreover, as exemplified by the prior inconsistent findings, collecting self-reported data from a sufficiently large cohort of relatives to accurately determine associations between rare exposures and outcomes remains a significant challenge.

Second, while healthcare registers in Finland and Sweden provide accurate admission and discharge dates, these dates do not reflect the actual onset of psychiatric disorders but rather diagnosis dates. Consequently, determining the temporal sequence of events is not entirely straightforward. Furthermore, patients with assault-related injuries may have a higher likelihood of referral to psychiatric services compared to the general population, which could lead to inflated rates of diagnosed psychiatric disorders in this group. To mitigate the risks for such biases, we initiated the follow-up period up to two years after the initial victimisation event in complementary sensitivity tests and found that the results were commensurate with the main findings. This time lag strengthens the argument that the associations are unlikely due to reverse causation bias or solely attributable to earlier referrals to psychiatric services. In addition, we examined premature mortality as outcome, as it is to a lesser extent affected by referral practices and found that it was similarly associated with earlier victimisation experience.

Third, while the sibling-comparison design is effective in controlling for shared unmeasured familial confounders, it does not address the impact of non-shared confounders (20). For this reason, some findings could be explained by residual genetic confounders that are not shared between siblings. However, we note that the observed within-family associations remain substantial. We also measure a number a previously suggested non-genetic confounders not shared between siblings, but observed in our data. Adjustments for such factors, such as low family income and having a single mother at birth, also increase the credibility of our findings. It is nevertheless important to note that sibling-comparison estimates may underestimate causal effects in the presence of misclassification bias, shared mediators, and sibling carryover effects (32,20). Future studies may benefit from triangulating our findings with alternative genetically informative research designs (33).

The generalisability of our findings is supported by two findings. First, rates of psychiatric disorders are similar across Western European countries (34), and self-reported violent victimisation rates in Finland and Sweden were closely aligned with the global average during the period 2003-2004 (2.2%-3.5% vs. 3.1%) when the last international comparison was conducted (35). Second, the country-specific associations in the present study were not systematically different from one another.

Our findings underline the importance of routinely considering how to prevent and manage victimisation risk in mental health services, and also the role of general hospital and emergency medicine to address mental health and re-victimisation. This will entail assessing risks in a structured and consistent way, which will enable stratification, allocation of additional resources to individuals at elevated risks, and potentially collaborative risk management with patients. Furthermore, research exploring the optimisation of trauma-based interventions could potentially reduce risks of post-victimisation risks of common psychiatric disorders and premature mortality. This includes investigations into tailoring therapies to specific populations and settings and evaluating the long-term effectiveness of existing approaches. Public health strategies can further broaden the scope beyond individual interventions, fostering closer collaboration with social services and criminal justice, and advocating for public policies targeting systemic factors like alcohol and substance use, which contribute to wider societal victimisation risks.

In summary, in this large cohort study, we found that individuals who experienced violent victimisation were at least twice as likely as their unaffected siblings to be diagnosed with common psychiatric disorders, to engage in suicidal behaviours, and to die prematurely. These risks remained elevated over two years post-victimisation, suggesting that mental health services should review how they assess and manage subsequent risks of adverse outcomes.

## Data Availability

Data may be obtained from a third party and are not publicly available. Finnish and Swedish privacy laws prohibit us from making individual-level data publicly available. Researchers who are interested in replicating our work using individual-level data can seek access via Findata, Statistics Sweden and the Swedish National Board for Health and Welfare. For more information, see https://findata.fi/en and https://www.scb.se/en/services/ordering-data-and-statistics/ordering-microdata/.

## Supporting information

**eTable 1.** STROBE statement

**eTable 2.** ICD diagnostic codes

**eText 1.** Diagnostic validity

**eTable 3.** Measured confounders

**eText 2.** Additional sensitivity tests

**eTable 4.** Baseline demographic characteristics in Finland

**eTable 5.** Baseline demographic characteristics in Sweden

**eTable 6.** Person-time at risk, number of individuals with the outcomes, and incident rates per 1000 person-years for common psychiatric disorders and suicidal behaviours, stratified across individuals exposed to violent victimisation in Finland and Sweden

**eTable 7.** Interaction terms between violent victimisation and pre-existing psychiatric disorders and self-harm on subsequent risk for common psychiatric disorders and suicidal behaviours in Finland and Sweden

**eFigure 1.** Country-specific within-family associations between violent victimisation and subsequent common psychiatric disorders and suicidal behaviours in Finland and Sweden

**eFigure 2.** Cohort-specific within-family associations between violent victimisation and subsequent common psychiatric disorders and suicidal behaviours in Finland and Sweden

**eFigure 3.** Age-specific within-family associations between violent victimisation and subsequent common psychiatric disorders and suicidal behaviours in Finland and Sweden

**eFigure 4.** Sex-specific within-family associations between violent victimisation and subsequent common psychiatric disorders and suicidal behaviours in Finland and Sweden

**eFigure 5.** Within-family associations between violent victimisation and subsequent common psychiatric disorders and suicidal behaviours in Finland and Sweden with varying washout periods (e.g., 1 month, 6 months, 12 months, and 24 months)

**eFigure 6.** Within-family associations between violent victimisation and subsequent common psychiatric disorders and suicidal behaviours in Finland and Sweden and adjusted for additional indicators for parental socioeconomic status (SES)

**eFigure 7.** Within-family associations between violent victimisation and subsequent common psychiatric disorders and suicidal behaviours (at least two diagnoses on separate occasions) in Finland and Sweden

**eFigure 8.** Within-family associations between violent victimisation and subsequent common psychiatric disorders and suicidal behaviours in Finland and Sweden stratified across the entire period and the most recent period (2006-2020)

